# Systematic Review and Meta-Analysis Identifies Significant Relationships Between Anxiety Disorders and Daytime Lower Urinary Tract Symptoms

**DOI:** 10.1101/2020.08.27.20183467

**Authors:** Behrang Mahjani, Lotta Renström Koskela, Anita Batuure, Christina Gustavsson Mahjani, Magdalena Janecka, Christina M. Hultman, Abraham Reichenberg, Joseph D. Buxbaum, Olof Akre, Dorothy E. Grice

**Affiliations:** Department of Psychiatry, Icahn School of Medicine at Mount Sinai, New York, NY, USA; Seaver Autism Center for Research and Treatment, Icahn School of Medicine at Mount Sinai, New York, NY, USA; Department of Medical Epidemiology and Biostatistics, Karolinska Institutet, Stockholm, Sweden; Department of Genetics and Genomic Sciences, Icahn School of Medicine at Mount Sinai, New York, NY, USA; Department of Neuroscience, Icahn School of Medicine at Mount Sinai, New York, NY, USA; The Mindich Child Health and Development Institute, Icahn School of Medicine at Mount Sinai, New York, NY, USA; Division of Tics, Obsessive-Compulsive Disorder (OCD) and Related Disorders, Icahn School of Medicine at Mount Sinai, New York, NY, USA; Friedman Brain Institute, Icahn School of Medicine at Mount Sinai. New York, NY, USA; Department of Molecular Medicine and Surgery, Karolinska Institutet, Stockholm, Sweden; Department of Pelvic Cancer, Karolinska Institutet, Stockholm, Sweden

**Author notes:** **Correspondence:** Dorothy E. Grice, M.D., 1425 Madison Avenue, New York, NY 10029. Contributed equally (Anita Batuure, Christina Gustavsson Mahjani).

**Keywords:** anxiety disorders, obsessive-compulsive disorder, lower urinary tract symptoms, overactive bladder, interstitial cystitis

## Abstract

**Background:** Clinically, lower urinary tract symptoms (LUTS), such as voiding symptoms, overactive bladder, and interstitial cystitis, are associated with anxiety disorders. However, the existing evidence for the overlap of these conditions has not been systematically evaluated. The aim of this study was to examine this relationship.

**Methods:** We conducted a systematic review and meta-analysis to examine the relationship between LUTS and anxiety. We searched for articles published from January 1990 to July 2019 in PubMed, CENTRAL, PsycINFO, and Google Scholar. Outcomes were anxiety-related disorders and symptoms “clinical anxiety”) and subtypes of LUTS. We performed random-effect meta-analyses, and inspected funnel plots and applied the Egger’s test to evaluate publication bias. We followed PRISMA guidelines and recorded our protocol on PROSPERO (ID=CRD42019118607).

**Results:** We identified 814 articles, of which 94 fulfilled inclusion criteria and 23 had sufficient data for meta-analysis. The odds ratio for clinical anxiety among individuals with LUTS was 2.87 (95% CI: 2.38,3.46, P < 0.001). Very few studies looked at LUTS among individuals ascertained for clinical anxiety, but there appear to be significantly elevated odds ratio here as well. A large I^2^ value suggests high heterogeneity between studies.

**Conclusions:** The results demonstrate a clinically significant association between clinical anxiety and LUTS. There were limited data on youth, which should motivate further study in this area. Understanding the co-occurrence of these conditions will lead to better prevention and interventions to ameliorate the progression of the symptoms and to improve quality of life.

## 1. Introduction

Anxiety disorders are among the most common psychiatric conditions, with lifetime prevalences as high as 33.7% (1). Although it is not uncommon to experience anxiety symptoms over the course of life, when the anxiety persists and impairs daily function, criteria for a clinical diagnosis may be met (2). Lower urinary tract symptoms (LUTS) are related to dysfunction of the lower urinary system, including the bladder, prostate, and urethra. LUTS can present across the life cycle in a variety of ways: through issues with storage (urgency, frequency, or incontinence), voiding, post micturition (Figure 1), as well as overactive bladder syndrome (OAB) and interstitial cystitis (IC). The population frequency of LUTS in adults is between 20-30% and varies due to definitions of LUTS (3, 4).LUTS can onset in childhood, with the prevalence increasing over time, and a secondary increase related to consequences of the aging process in older adults. Here, we focus on daytime LUTS in children, adolescents and younger adults and avoid LUTS related to aging (5). Specific LUTS symptoms, such as increased urinary frequency, pressure, urgency, and/or pain when the bladder fills, can have significant and direct effects on overall quality of life (6). The experience of LUTS in youth can put them at risk for social isolation, teasing, and other challenges, and they may experience low self-esteem or distress (7). Studies in children and adults with LUTS indicate that those with LUTS are more likely to also have anxiety symptoms, anxiety disorders, obsessive-compulsive disorder (OCD), and attentional problems (Table S1). Of note, a large longitudinal population-based cohort study argues that there is a bidirectional improvement when treating either LUTS or psychiatric symptoms (8). To date, no meta-analyses of associations between anxiety disorders and LUTS have been published. We hypothesis that there is a strong association between anxiety disorders, anxiety symptoms, and LUTS. In order to shed light on these associations, we conducted a systematic review and meta-analysis, examining studies in both children and adults.

**Figure 1.**
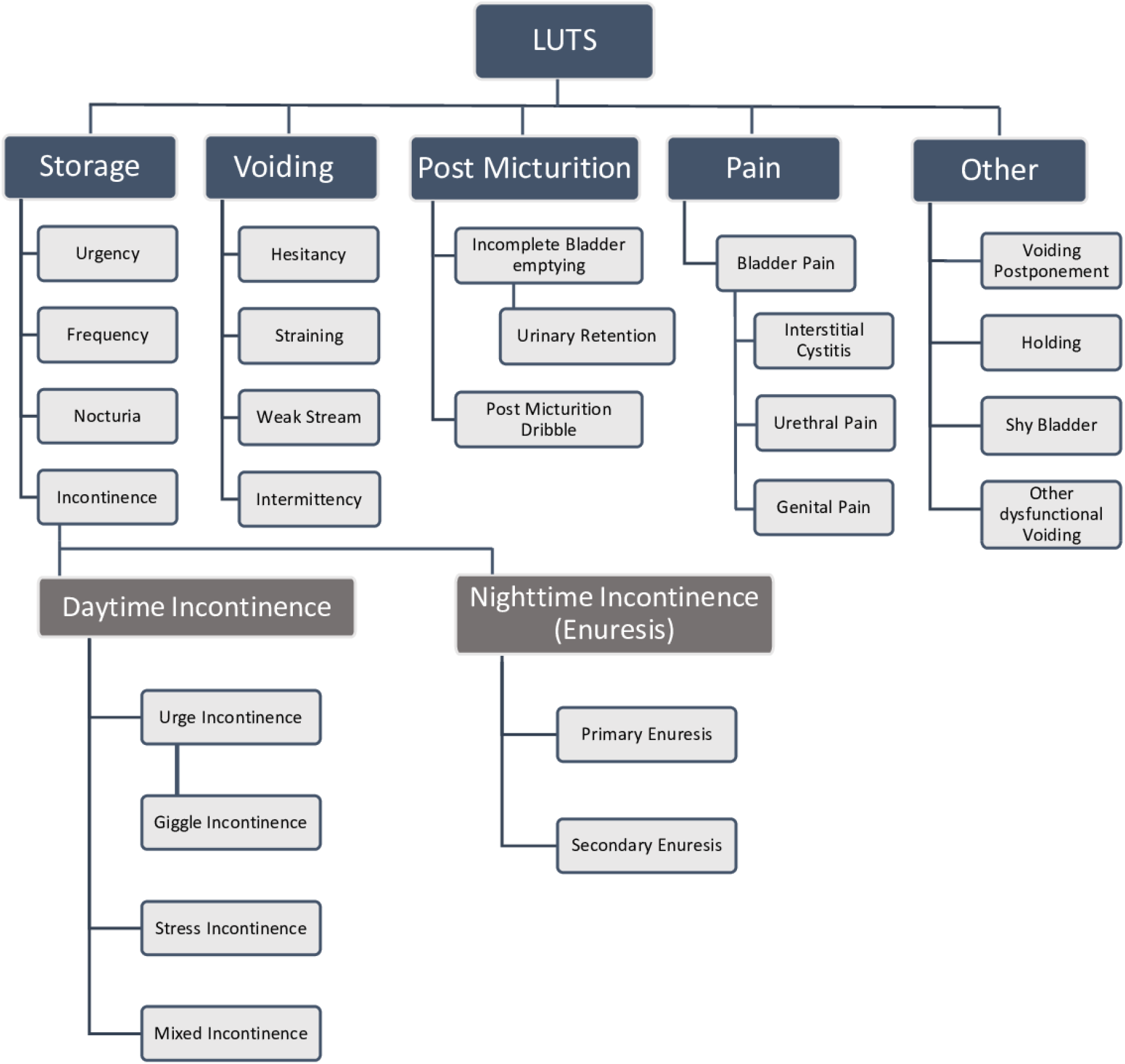
Subcategories of lower urinary tract symptoms (LUTS).

## 2. Material and methods

### 2.1 Search strategy and selection criteria

We recorded our approach using the PRISMA protocol (9) (Supplement 1) in PROSPERO (10) (ID=CRD42019118607). Figure 2 outlines the steps used to search the literature for qualifying articles. We summarize the overall approach here and provide greater detail further in the Methods. (1) *Identification:* We identified all articles with results in humans that included our search terms referencing anxiety disorders or LUTS in the title OR abstract, and anxiety disorders AND LUTS in the main text (Supplement 2); (2) *Screening:* At least two authors (BM, MJ, AB) independently screened all identified articles for inclusion/exclusion criteria, based on titles and abstracts, and twenty articles were randomly chosen and re-screened by a third author (DG); (3) *Eligibility:* At least two authors (CM, AB, BM) independently reviewed the full text of each selected article and applied the exclusion criteria. Excluded articles and conflicts were reviewed by a different author and, again, 20 articles were randomly chosen and re-screened by DG. We excluded studies with sample sizes of less than 20 individuals, samples where all individuals were older than 50 years old (due to the impact of aging on urinary symptoms), articles reporting only nocturnal enuresis or when it was not possible to separate the study results for daytime versus nighttime wetting. We also excluded treatment trials, because it was not possible to discern if clinical symptoms were secondary to the intervention or independent of it.

**Figure 2.**
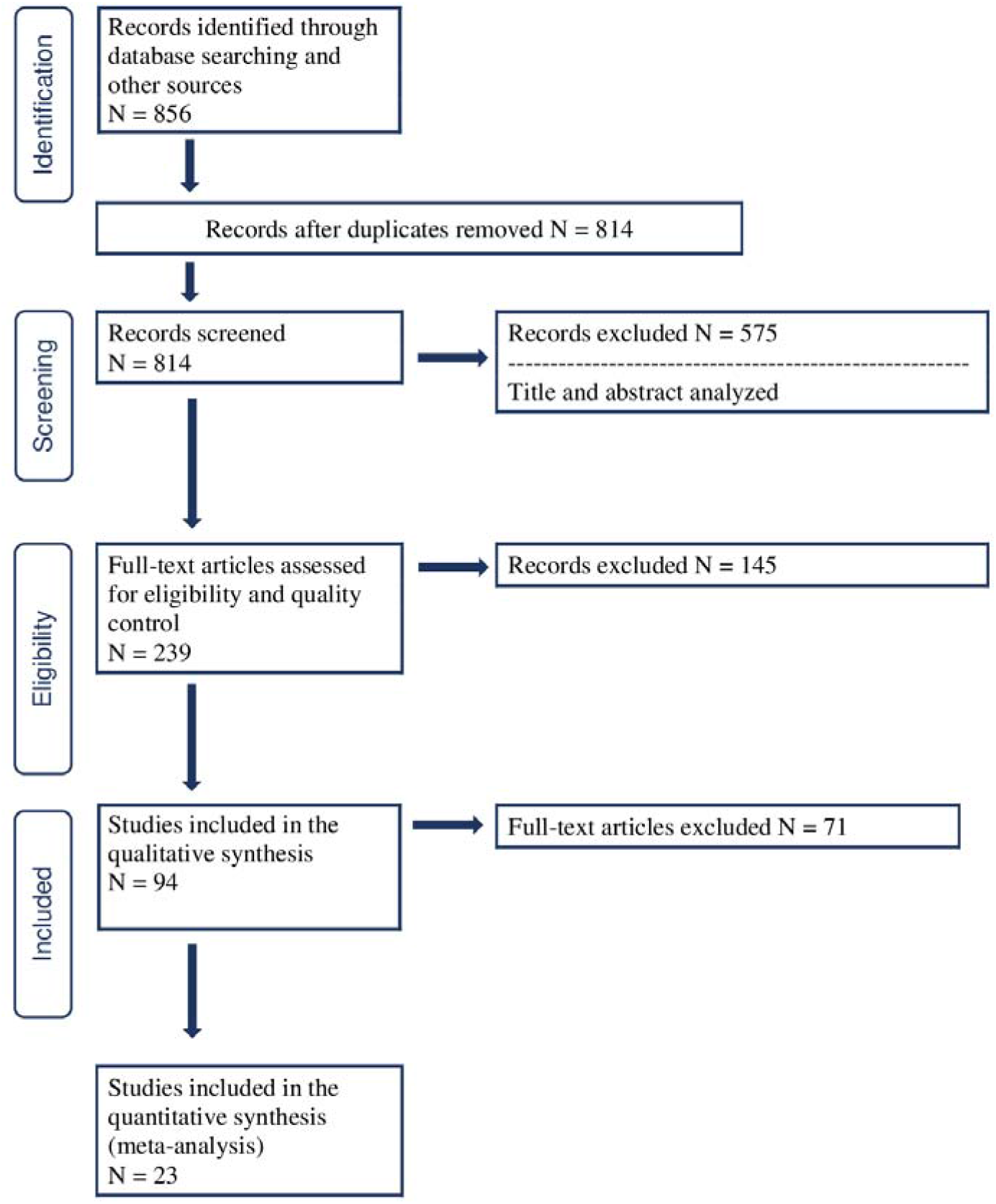
PRISMA details.

We searched systematically for articles in PubMed, CENTRAL, PsycINFO, and Google Scholar (specific search terms in Supplement 2). We used Google Scholar to find any articles that might have been missed by the searches via PubMed, CENTRAL, and PsycINFO. We performed 12 searches in Google Scholar using different keywords (details in Supplement 2). For each search, we screened the first 100 articles and identified only a single article that was not found previously (details in Supplement 2). We also examined all references from relevant review articles to find any eligible articles to complement the database searches.

Observational studies were included in the review (cohort studies, case-control studies, and cross-sectional studies). We did not include ecological, treatment, or case series studies. We considered peer-reviewed, published studies (including studies published online ahead of print or online only) in English (or translated into English) from January 1, 1990, to July 15, 2019. We excluded all studies published as editorials or commentaries. We did not seek data from any unpublished studies.

For each manuscript, we checked the quality of study design, the power of the study, and the use of appropriate statistical techniques. Each article review was carried out by at least two individuals (MJ, BM, AB, CM) who independently utilized a checklist of clinical parameters, methodology and results to determine eligibility for inclusion. Any conflicts in opinion were discussed until a consensus was reached. A second-tier review was performed by experts in child psychiatry, urology, and statistics (DG, LK, OA, JB) who assessed study summaries and checklists, and final inclusion was determined by expert consensus.

### 2.2 Psychiatric Outcomes

Anxiety disorders cover a broad spectrum of conditions. Generalized anxiety disorder (GAD), social anxiety disorder, separation anxiety, panic disorder and specific phobias occur across the lifespan, with significant rates in children, adolescents and adults (3,4). Selective mutism is largely described in younger school age children but it can continue through childhood, particularly in the absence of treatment. Although OCD and posttraumatic stress disorder (PTSD) were moved from the anxiety disorder section of DSM IV (Diagnostic and Statistical Manual of Mental Disorders, fourth edition) to separate sections in DSM 5 (Diagnostic and Statistical Manual of Mental Disorders, fifth edition), albeit with minimal changes in diagnostic criteria, anxiety symptoms are also at the core of these conditions (5,6). Pediatric acute-onset neuropsychiatric syndrome (PANS) is characterized by an abrupt onset of obsessive-compulsive symptoms or eating restrictions. When associated with streptococcal infection, the condition is subtyped as pediatric autoimmune neuropsychiatric disorder associated with streptococcal infections (PANDAS) (12–15). In the trauma- and stressor-related section of DSM 5, the diagnosis of PTSD requires exposure to a life-threatening trauma, which can occur at any time across the life cycle. The psychiatric sequelae of PTSD are manifest by severe anxiety symptoms, albeit brought on an external exposure, unlike the anxiety disorders described above. In this study, we used the following psychiatric outcomes: GAD, social anxiety disorder, separation anxiety, adjustment disorder, selective mutism, panic disorder, agoraphobia, phobias, OCD and PTSD. We also included all possible cases of anxiety disorders based on elevated anxiety symptoms, even where the specific reference to ICD/DSM was missing, provided the study participants obtained a clinically significant result on standardized questionnaires/measures. Standardized questionnaires, such as Hospital Anxiety and Depression Scale (HADS), Zung Self-Rating Anxiety Scale (SAS), and General Anxiety Disorder 7-item (GAD-7) do not include all of the diagnostic criteria of anxiety disorders and are primarily designed to measure anxiety symptoms and detect possible cases. In this work, we defined *clinical anxiety* as having an anxiety related disorder or elevated anxiety symptoms based on standardized questionnaires.

### 2.3 Urological Outcomes

Urinary incontinence can be divided into subgroups and in many publications authors make a distinction between enuresis/nocturnal enuresis and daytime urinary incontinence (Figure 1). However, in both ICD-10 (International Classification of Diseases, Tenth Revision) and DSM 5, enuresis and daytime incontinence are coded as one disorder, without differentiating between subtypes of incontinence (11). Enuresis is defined as any intermittent incontinence while sleeping (at night or during daytime naps). Daytime incontinence includes: urge incontinence, stress incontinence and mixed incontinence (Figure 1) (12).

OAB and IC are storage LUTS with considerable symptomatic overlaps. Individuals with OAB can experience frequency, urgency, and nocturia. OAB can occur at any age, although the prevalence of the different symptoms varies across age groups (13). IC or painful bladder syndrome is characterized by lower abdominal pain and pressure. IC has been rarely reported among children (14). IC and OAB have similar symptoms, making them difficult to differentiate. Pelvic pain is a common symptom of IC but generally not present in patients with OAB (15).

In this study, we used the following urological outcomes: overactive bladder, bladder pain syndrome, interstitial cystitis, urinary storage symptoms, urinary voiding symptoms, and daytime urinary incontinence. We excluded LUTS associated with physical trauma, cancer, sexually-transmitted infection, congenital malformation of the urinary tract, post-operative disorders, those related primarily to pregnancy, dysfunction secondary to aging, and medications that promote urination/water retention. We excluded outcomes associated with kidney malfunction, unless they arose as a consequence of a lower urinary tract infection. We included LUTS due to infection, as regulation of the immune system could be a potential common factor linking urinary and psychiatric symptoms.

### 2.4 Statistical analysis

For the meta-analysis, we extracted the number of exposed/unexposed individuals with LUTS, or clinical anxiety from the published articles to calculate the odds ratio. We excluded articles that did not provide the data required for meta-analysis. If multiple articles analyzed the same cohort, then we chose only one article. The screening was completed using DistillerSR (Evidence Partners, Ottawa, Canada).

Utilizing data from eligible studies, we performed meta-analyses, employing random-effects models using reciprocal of the estimated variance, allowing for combining effect estimates without access to raw data. We reported two measures of heterogeneity, the Cochran’s Q test, and the I^2^ index. To evaluate publication bias, we visually inspected funnel plots and applied Egger’s test (16). We used *metafor* package in R for the meta-analysis (17).

We did two sensitivity analyses to shed light on the heterogeneity of the results. (1) We dissected the outcomes for clinical anxiety into anxiety disorders based on ICD codes, anxiety symptoms based on HADS, and anxiety symptoms based on other scales; (2) We dissected the outcomes for LUTS into IC, OAB, urinary incontinence, and other LUTS.

## 3. Results

We identified 814 articles published from January 1, 1990 to July 15, 2019 that met the inclusion criteria (Figure 2, *Identification*). Following review of titles and abstracts, and then full texts, 94 articles were included in qualitative and quantitative synthesis. The characteristics of these articles are summarized in Table S1. 23 articles were included in the meta-analysis, and are summarized in Table S2.

There were two striking initial findings. First, there was only one study in pediatric clinical anxiety with sufficient data for inclusion in the meta-analysis (18). Second, the overwhelming majority of studies suitable for inclusion in the meta-analysis involved individuals first ascertained with a LUTS diagnosis, who were then evaluated for anxiety phenotypes (n=21). Only 2 studies suitable for meta-analysis examined individuals ascertained for clinical anxiety who were then assessed for LUTS (8, 19). Hence, beyond a perfunctory understanding of these relationships, there are clear gaps in the literature.

### 3.1 Clinical anxiety among individuals diagnosed with LUTS

The largest group of qualifying studies were those examining anxiety disorders, or their proxy, in individuals ascertained with LUTS, 21 of which had sufficient data for meta-analysis. Two studies included several anxiety diagnoses and we selected the larger clinical category. Using data from the Avon Longitudinal Study of Parents and Children (ALSPAC), Joinson *et al*. (18) included separation anxiety, social fears, and general anxiety, and we chose general anxiety because it had the largest number of subjects. Talati *et al*. (19) included both panic disorder and social anxiety disorder, and we chose the panic disorder category, which was larger. Using a random effect model, the odds ratio for clinical anxiety among cases with LUTS was 2.87 (95% CI: 2.38,3.46, P < 0.001; Figure 3). When we excluded PTSD from the core anxiety group, the odds ratio for the association between clinical anxiety and LUTS was 3.01 (95% CI: 2.44,3.71, P < 0.001; Figure S1).

**Figure 3.**
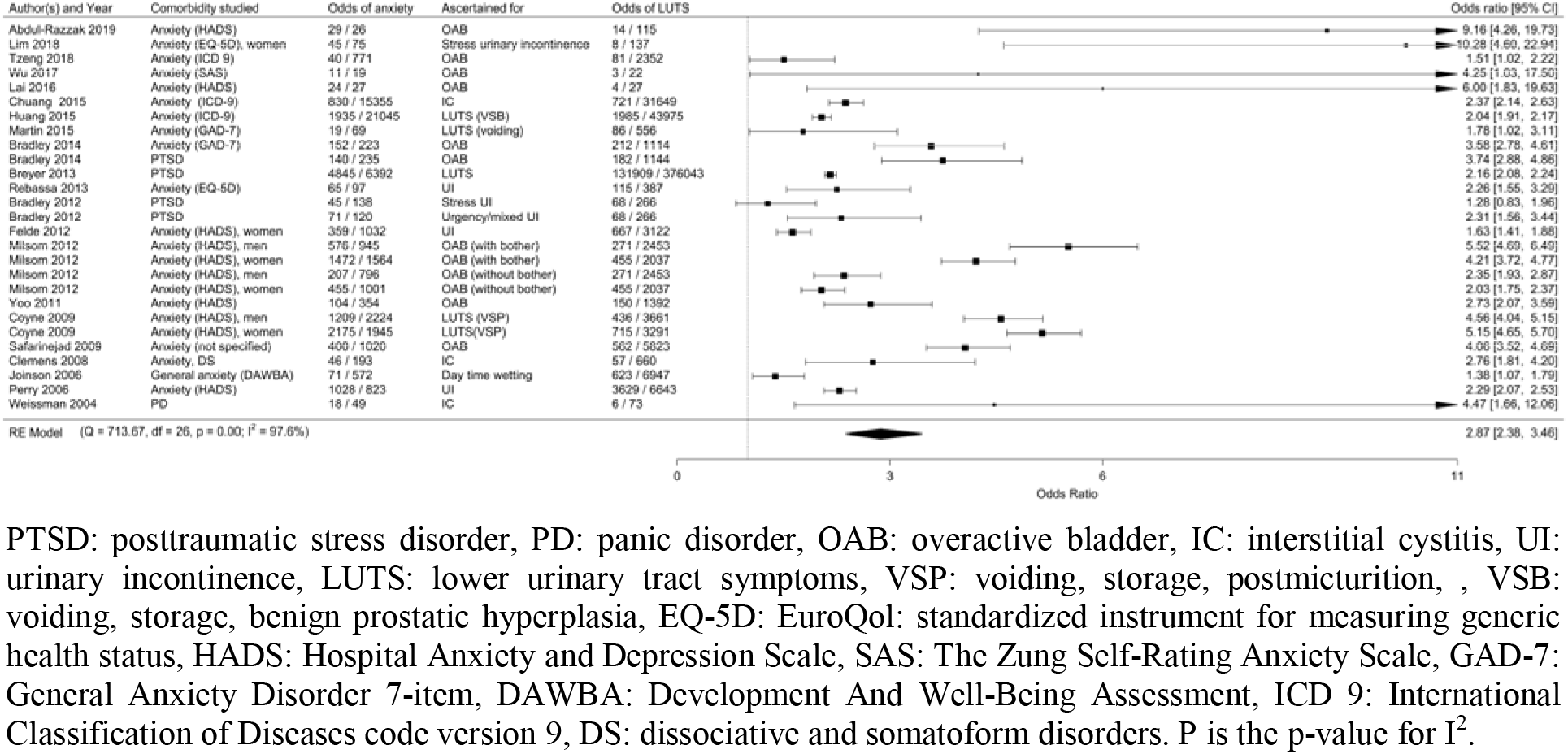
Forest plot for clinical anxiety among individuals with LUTS.

Because the rates of LUTS differs between men and women (20), we also examined the qualifying articles for differential risk as a function of sex. Few studies qualified for this sub-analysis. Among these studies, the odds ratio for clinical anxiety among women with LUTS was 3.14 (95% CI: 1.85,5.3, P < 0.001) and for men it was 3.52 (95% CI: 2.34,5.30, P < 0.001; Figure 4).

**Figure 4.**
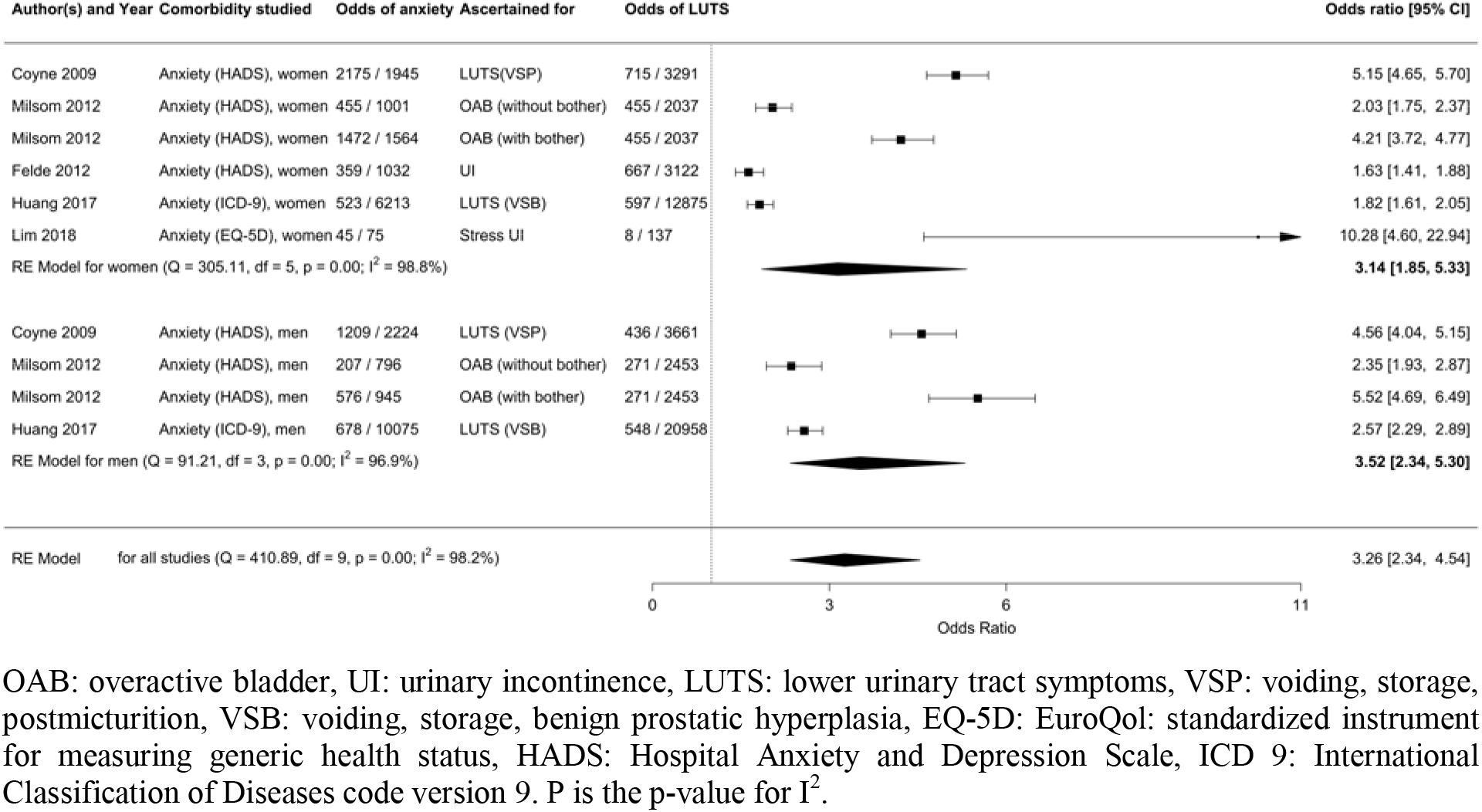
Forest plot for clinical anxiety among individuals with LUTS, comparing men and women.

Of the 94 articles that investigated an association between clinical anxiety and LUTS, 16 articles focused on children (18, 21–35) and only one article had the required information for meta-analysis (18). Hence, we could not perform a child-specific meta-analysis. However, our qualitative review of these studies shows that between 1-20% of children with LUTS had clinical anxiety (18, 21, 28, 29, 31–34), and 19-46% had at least one psychiatric comorbidity, with attention-deficit/hyperactivity disorder (ADHD) having the highest rate (21, 23, 28, 31, 33, 34).

### 3.2 LUTS in individuals diagnosed with clinical anxiety

Only 2 articles (8, 19) had sufficient data for meta-analysis of LUTS in individuals ascertained with clinical anxiety. The odds ratio was 2.87 (95% CI: 1.07,7.74, P =0.037; Figure S2), similar to the studies in which individuals were ascertained for LUTS.

### 3.3 Bidirectional studies

In a study of the association between anxiety disorders (anxiety states, phobic disorders, obsessive-compulsive disorders, and adjustment disorder with anxiety) and LUTS, after controlling for age, sex, and medical comorbidities, individuals with LUTS were 2.12 (95% CI: 1.95-2.30) times more likely to develop anxiety disorders, and individuals with anxiety disorders were 2.01 (95%CI: 1.88-2.14) times more likely to develop LUTS (8). In another study, urinary incontinence and frequency were predictors of incident cases of anxiety (measured using HADS), and anxiety was a predictor of incident cases of urinary incontinence (36).

### 3.4 Association of LUTS and OCD

We were also interested in the specific relationship between OCD and LUTS. We found 12 articles discussing this association (19, 22, 25, 27, 30, 32, 35, 37–41), of which 6 focused on children (22, 25, 27, 30, 32, 35). Only one of these 12 articles was suitable for meta-analysis although there did appear to be an association between OCD and LUTS based on the content. Review of smaller studies of PANS/PANDAS suggests that urinary urgency, frequency and enuresis are prominent clinical symptoms in children diagnosed with OCD that is categorized as PANS/PANDAS (22, 25, 27, 30, 35).

### 3.5 Analysis of heterogeneity

We analyzed the heterogeneity of the studies for clinical anxiety among individuals diagnosed with LUTS. Visual inspection of the funnel plot (Figure 5) and the large value of I^2^ and Cochran’s Q test suggest a high heterogeneity between studies. The Egger test was not statistically significant (p-value=0.16), rejecting the null hypothesis that the funnel plot is asymmetrical. Nevertheless, the power of this method to detect bias is low with small numbers of studies. By visual inspection of the funnel plot (Figure 5), we observed an asymmetrical pattern mainly driven by five studies with large effect sizes but small sample sizes. Studies with small samples and negative effects were absent in our meta-analysis. Funnel plot with the Trim and Fill method (Figure S3) suggests at least four studies with small samples and negative effects were missing for a symmetrical funnel (42). However, after adjusting with Trim and Fill method, I^2^ did not decrease (I^2^= 97.86) The majority of the eligible studies reported at least one domain of psychiatric disorders or urinary symptoms, therefore most negative studies may be unpublished. There were between-study variations in the methods of assessment for clinical anxiety and LUTS, which may have contributed to the heterogeneity of pooled estimates (see sensitivity analysis).

**Figure 5.**
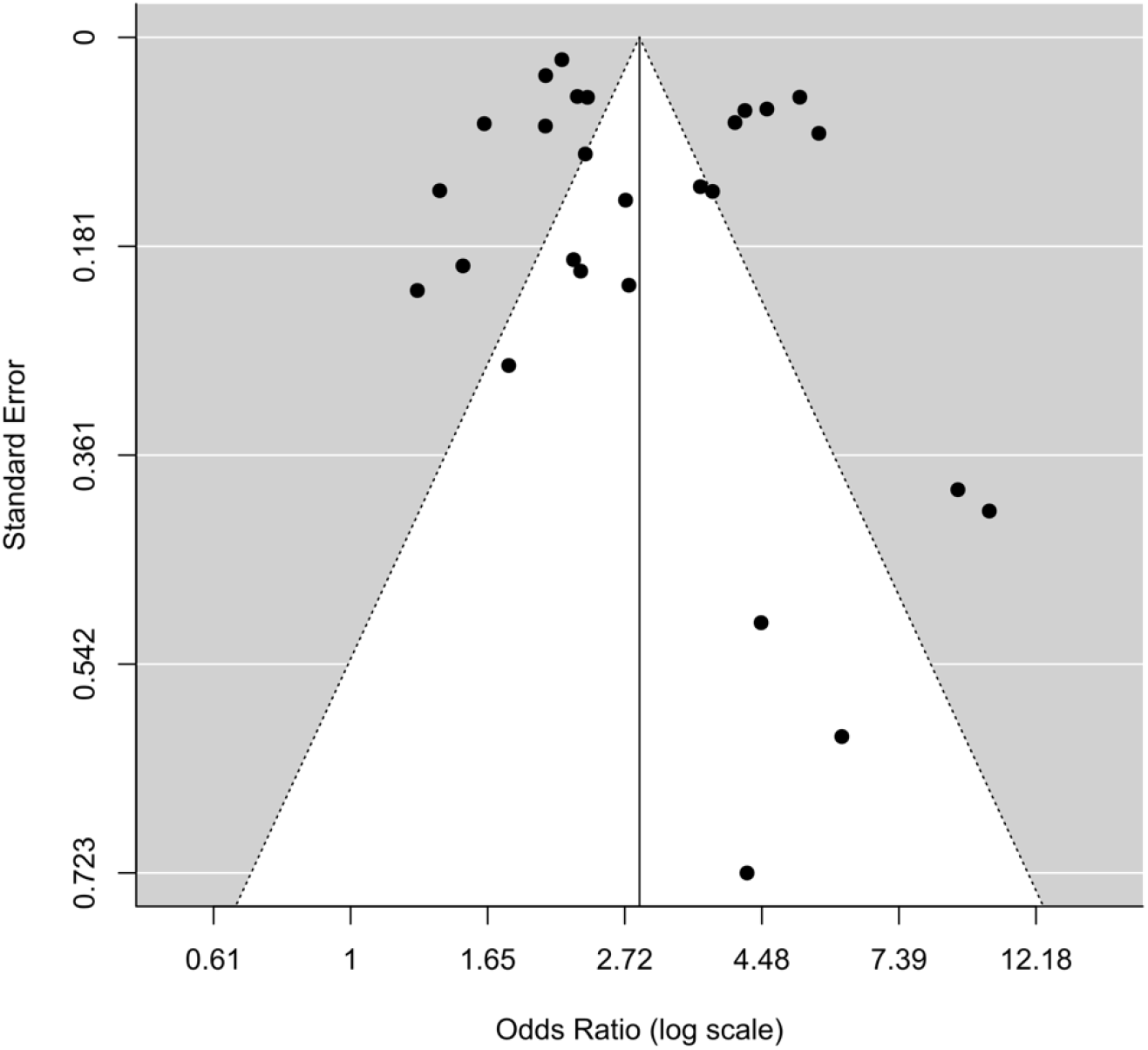
Funnel Plot

### 3.6 Sensitivity analysis

We performed two sensitivity analyses. For the first analysis, we categorized clinical anxiety outcomes into three categories (1) anxiety disorders based on ICD code, (2) anxiety symptoms based on the HADS scale, and (3) anxiety symptoms based on other scales. We did not include PTSD in this analysis. For the HADS group, we estimated OR=3.41 [95% CI: 2.53,4.60, P<0.001], Q=356.01, I^2^=97.4%; for the ICD-9 group OR=2.07 [95% CI: 1.72,2.49, P=0.01], Q=9.16, I^2^=84.6%; and for the other scales OR=3.03 [95% CI: 2.08,4.41, P<0.001], Q=67.79, I^2^=88.9% (Figure S4). The ICD group had the lowest heterogeneity in comparison to the two other groups.

For the second analysis, we categorized LUTS outcomes into four categories (1) IC, (2) OAB, (3) urinary incontinence, and (4) other LUTS. For the IC group, we estimated OR=2.41 [95% CI: 2.18,2.66, P=0.37], Q=1.98, I^2^=0%; for the OAB group OR=3.39 [95% CI: 2.53,4.55, P=<0.001], Q=136.19, I^2^=94.1%; for the urinary incontinence group OR=2.82 [95% CI: 1.39,5.71, P=0.0041], Q=29.6, I^2^=97.9%%; and for the other LUTS group OR=2.65 [95% CI: 1.57,4.47, P=0.0015], Q=324.03, I^2^=98.8% (Figure S5). The IC group had the lowest heterogeneity; however, the estimate of I^2^ was not significant due to the small sample size.

## 4. Discussion

The results of this systematic review and meta-analysis demonstrate reliable associations between clinical anxiety and LUTS. Risk for clinical anxiety amongst individuals with a LUTS diagnosis was 2.87 (95% CI: 2.38,3.46), or 3.01 (95% CI: 2.44,3.71) when excluding PTSD. The odds ratio for clinical anxiety among women with LUTS was slightly lower than men (3.14 vs. 3.52), although the number of included studies was small and the confidence intervals were overlapping. When examining the studies in which individuals were ascertained for clinical anxiety and then assessed for LUTS, findings were similar, with an odds ratio of 2.87 (CI: 1.07,7.74). However, this result should be interpreted with caution since the sample size for this analysis was small (n=2), which is reflected in the wide confidence interval.

In all analyses, we observed a large I^2^ index and Cochran’s Q test, suggesting a high heterogeneity. The heterogeneity may be related to bias from publication, reporting, and selection, as supported by many studies plotting outside the funnel outline in the funnel plot (Figure 5). We analyzed the sensitivity of the results to the definition of the outcomes for clinical anxiety and LUTS. The results from sensitivity analyses suggested that one of the leading causes of the high degree of heterogeneity in the results was the inhomogeneous definition of anxiety outcomes. The estimate of I^2^ was the lowest when we categorized clinical anxiety based on ICD codes, which was the most well-defined category. Although we estimated different values of I^2^ for different dissections of outcomes in sensitivity analyses, OR was larger than 2 for all the analyses, and confidence intervals were never overlapping 1. Another interesting observation in the sensitivity analysis was that OAB had the highest OR among other LUTS categories.

In sum, there is compelling evidence for a strong association between clinical anxiety and LUTS and it appears to be both bidirectional and clinically significant. Multiple studies were cross-sectional and causality of the association in either direction could not be accurately determined using meta-analysis. However, there were supports in the literature for a bidirectional association. A large longitudinal cohort study provides evidence for a bidirectional relationship between LUTS and anxiety disorders (8) and a second longitudinal study showed that anxiety (measured using HADS) was both a risk factor for, and a consequence of, urinary incontinence (36).

It was interesting to note that the association of anxiety disorders with LUTS appears to cross diagnostic categories. Previously, Weissman *et al*. posited a syndrome manifested by interstitial cystitis and panic disorder (43). However, our study shows that multiple anxiety disorder symptoms and multiple lower urinary tract symptoms show a similar relationship (Figure 3). Among psychiatric diagnoses, this relationship also extended to PTSD which is characterized by prominent anxiety-related symptoms although not classified in the DSM 5 anxiety disorder section due to its alternate phenomenology. We examined the result with and without PTSD because of the prominent anxiety symptoms that manifest in this disorder.

Our analyses also highlight that there are very significant gaps in the literature. Firstly, studies of LUTS in individuals ascertained for an anxiety diagnosis or clinical anxiety symptoms are rare. Studies in mental health settings are needed to elaborate on these associations so they can be more fully understood. Similarly, studies using national registers, where ascertainment and access issues are less confounding, could help us understand these associations, particularly since data are collected prospectively across a range of health care visits, related to both mental health and medical conditions, and additional demographic information can enrich our understanding of these relationships. Secondly, diagnostic criteria and definitions are not always consistent or clear in the existing studies. For example, there were few studies that made use of defined diagnoses and criteria. Some studies used severity measures and others used approaches that would include or exclude subclinical diagnoses in the psychiatric phenotypes. Therefore, we pooled the outcomes based on elevated levels of anxiety symptoms rather than formal anxiety diagnoses alone and defined the outcome as clinical anxiety. It is apparent that in order to understand relationships with specific diagnoses, more studies are needed. Finally, and most strikingly, is the absence of studies of pediatric anxiety disorders. This is important to address for several reasons: (i) childhood LUTS is impairing and is predictive of adult LUTS, hence treating LUTS (including associated anxiety) may have both immediate and long-term impact; (ii) rates of LUTS increase with age, therefore the study of pediatric age individuals may reveal differing findings and risks; furthermore, (iii) childhood-onset anxiety disorders may differ from adult-onset anxiety disorders so, again, it may be inappropriate to generalize from adult studies to studies of youth. Given the strong findings in adults, we would argue that studies in children, as well as longitudinal studies, are critically needed.

In summary, our systematic review and meta-analysis of the published literature indicate a significant association across multiple anxiety diagnoses, anxiety symptoms, and multiple lower urinary tract symptoms, regardless of the definition of clinical anxiety or LUTS. Careful evaluation for these co-morbid conditions represents an important aspect of clinical care, with referral to appropriate specialists, if indicated. This strong association should motivate addressing the significant knowledge gaps in this area, especially the dearth of data on children. Understanding the causes of the observed relationships, be they biological or behavioral, is an important topic for future study.

## Data Availability

data available upon request

## Author Contributions

Mahjani had full access to all the data in the study and takes responsibility for the integrity of the data and the accuracy of the data analysis.

## Study concept and design

Akre, Batuure, Buxbaum, Grice, Gustavsson Mahjani, Janecka, Koskela, Mahjani

## Acquisition, analysis, or interpretation of data

Akre, Buxbaum, Grice, Janecka, Koskela, Mahjani

## Drafting of the manuscript

Batuure, Buxbaum, Grice, Gustavsson Mahjani, Janecka, Koskela, Mahjani

## Critical revision of the manuscript for important intellectual content

All authors.

## Statistical analysis

Mahjani

## Obtained funding

Grice

## Study supervision

Akre, Buxbaum, Grice, Koskela

## Availability of data

The data that support the findings of this study are available from the corresponding author upon reasonable request.

## Acknowledgments

This study was supported by a grant from the Friedman Brain Institute (DEG); the Beatrice and Samuel A. Seaver Foundation (DEG, MJ, JDB, BM) Icahn School of Medicine at Mount Sinai, New York, NY; the Mindworks Charitable Lead Trust (DEG); the Stanley Center for Psychiatric Research (DEG and JDB).

The sponsors of the study had no role in study design, data collection, data analysis, data interpretation, writing of the report, or in the decision to submit the paper for publication. The corresponding authors had full access to all data in the study and had final responsibility for the decision to submit for publication.

## Supporting Information

-Supplement 1:PRISMA checklist

-Supplement 2:Database Search Strategy

-Table S1. Characteristics of included studies (*n* = 94)

-Table S2. Characteristics of included studies in the meta-analysis (*n* = 23)

-Figure S1. Forest plot for clinical anxiety among individuals with LUTS, excluding the results for PTSD.

-Figure S2. Forest plot for LUTs among individuals with clinical anxiety.

-Figure S3. Funnel Plot, Trim and Fill method

-Figure S4. Sensitivity analysis of clinical anxiety outcomes

-Figure S5. Sensitivity analysis of LUTs outcomes

## References

1. Bandelow B, Michaelis S: Epidemiology of anxiety disorders in the 21st century. Dialogues Clin Neurosci 2015; 17:327–335

2. Keller MB, Lavori PW, Wunder J, et al.: Chronic Course of Anxiety Disorders in Children and Adolescents. J Am Acad Child Adolesc Psychiatry 1992; 31:595–599

3. Coyne KS, Sexton CC, Thompson CL, et al.: The prevalence of lower urinary tract symptoms (LUTS) in the USA, the UK and Sweden: Results from the epidemiology of LUTS (EpiLUTS) study. BJU Int 2009; 104:352–360

4. Soler R, Gomes CM, Averbeck MA, et al.: The prevalence of lower urinary tract symptoms (LUTS) in Brazil: Results from the epidemiology of LUTS (Brazil LUTS) study. Neurourol Urodyn 2018; 37:1356–1364

5. Boyle P, Robertson C, Mazzetta C, et al.: The prevalence of lower urinary tract symptoms in men and women in four centres. The UrEpik study. BJU Int 2003; 92:409–414

6. Milsom I, Kaplan SA, Coyne KS, et al.: Effect of bothersome overactive bladder symptoms on health-related quality of life, anxiety. Urology 2012;

7. Thibodeau BA, Metcalfe P, Koop P, et al.: Urinary incontinence and quality of life in children. J Pediatr Urol 2013; 9:78–83

8. Huang CLC, Wu MP, Ho CH, et al.: The bidirectional relationship between anxiety, depression, and lower urinary track symptoms: A nationwide population-based cohort study. J Psychosom Res 2017; 100:77–82

9. Moher D, Liberati A, Tetzlaff J, et al.: Preferred reporting items for systematic reviews and meta-analyses: the PRISMA statement. J Clin Epidemiol 2009; 62:1006–1012

10. PROSPERO International prospective register of systematic reviews 2020;

11. von Gontard A, Equit M: Comorbidity of ADHD and incontinence in children. Eur Child Adolesc Psychiatry 2014; 24:127–140

12. Austin PF, Bauer SB, Bower W, et al.: The standardization of terminology of lower urinary tract function in children and adolescents: Update report from the standardization committee of the International Children’s Continence Society. Neurourol Urodyn 2016; 35:471–481

13. McLennan MT: Interstitial cystitis: Epidemiology, pathophysiology, and clinical presentation. Obstet Gynecol Clin North Am 2014; 41:385–395

14. Close CE, Carr MC, Burns MW, et al.: Interstitial Cystitis in Children. J Urol 1996; 156:860–862

15. Macdiarmid SA, Sand PK: Diagnosis of interstitial cystitis/ painful bladder syndrome in patients with overactive bladder symptoms. Rev Urol 2007; 9:9–16

16. Schwarzer G, Carpenter JR, Rücker G: Meta-Analysis with R (Use R!). 1st ed. Springer, 2015

17. Viechtbauer W: Conducting meta-analyses in R with the metafor. J Stat Softw 2010; 36:1–48

18. Joinson C, Heron J, von Gontard A: Psychological Problems in Children With Daytime Wetting. Pediatrics 2006; 118:1985–1993

19. Talati A, Ponniah K, Strug LJ, et al.: Panic Disorder, Social Anxiety Disorder, and a Possible Medical Syndrome Previously Linked to Chromosome 13. Biol Psychiatry 2008; 63:594–601

20. Maserejian NN, Chen S, Chiu GR, et al.: Incidence of lower urinary tract symptoms in a population-based study of men and women. Urology 2013; 82:560–564

21. Zink S, Freitag CM, von Gontard A: Behavioral Comorbidity Differs in Subtypes of Enuresis and Urinary Incontinence. J Urol 2008; 179:295–298

22. Jaspers-Fayer F, Chan E, Ellwyn R, et al.: Prevalence of Acute-Onset Subtypes in Pediatric Obsessive-Compulsive Disorder. J Child Adolesc Psychopharmacol 2017; 27:332–341

23. Von Gontard A, Lettgen B, Olbing H, et al.: Behavioural problems in children with urge incontinence and voiding postponement: a comparison of a paediatric and child psychiatric sample. BJU Int 1998; 81:100–106

24. Von Gontard A, Moritz AM, Thome-Granz S, et al.: Abdominal pain symptoms are associated with anxiety and depression in young children. Acta Paediatr Int J Paediatr 2015; 104:1156–1163

25. Frankovich J, Thienemann M, Pearlstein J, et al.: Multidisciplinary Clinic Dedicated to Treating Youth with Pediatric Acute-Onset Neuropsychiatric Syndrome: Presenting Characteristics of the First 47 Consecutive Patients. J Child Adolesc Psychopharmacol 2015; 25:38–47

26. Filce HG, Lavergne LC: The impact of a 1-week residential program on anxiety in adolescents with incontinence: A quasi-experimental study. J Wound, Ostomy Cont Nurs 2013; 40:185–192

27. Bernstein GA, Victor AM, Pipal AJ, et al.: Comparison of Clinical Characteristics of Pediatric Autoimmune Neuropsychiatric Disorders Associated with Streptococcal Infections and Childhood Obsessive-Compulsive Disorder. J Child Adolesc Psychopharmacol 2010; 20:333–340

28. Wolfe-Christensen C, Veenstra AL, Kovacevic L, et al.: Psychosocial difficulties in children referred to pediatric urology: A closer look. Urology 2012; 80:907–913

29. Kuizenga-Wessel S, Koppen IJN, Vriesman MH, et al.: Attention Deficit Hyperactivity Disorder and Functional Defecation Disorders in Children. J Pediatr Gastroenterol Nutr 2018; 66:244–249

30. Swedo SE, Seidlitz J, Kovacevic M, et al.: Clinical Presentation of Pediatric Autoimmune Neuropsychiatric Disorders Associated with Streptococcal infections in Research and Community Settings. J Child Adolesc Psychopharmacol 2015; 25:26–30

31. Schast AP, Zderic SA, Richter M, et al.: Quantifying demographic, urological and behavioral characteristics of children with lower urinary tract symptoms. J Pediatr Urol 2008; 4:127–133

32. Özen MA, Mutluer T, Necef I, et al.: The overlooked association between lower urinary tract dysfunction and psychiatric disorders: a short screening test for clinical practice. J Pediatr Urol 2019;

33. Oliver JL, Campigotto MJ, Coplen DE, et al.: Psychosocial comorbidities and obesity are associated with lower urinary tract symptoms in children with voiding dysfunction. J Urol 2013; 190:1511–1515

34. Niemczyk J, Equit M, Rieck K, et al.: EEG measurement of emotion processing in children with daytime urinary incontinence. Z Kinder Jugendpsychiatr Psychother 2018; 46:336–341

35. Murphy M., Pichichero M.: Prospective identification and treatment of children with pediatric autoimmune neuropsychiatric disorder associated with group A streptococcal infection (PANDAS). Arch Pediatr Adolesc Med 2002; 156:356–361

36. Perry S, McGrother CW, Turner K: An investigation of the relationship between anxiety and depression and urge incontinence in women: Development of a psychological model. Br J Health Psychol 2006; 11:463–482

37. Hsiao SM, Liao SC, Chen CH, et al.: Psychometric assessment of female overactive bladder syndrome and antimuscarinics-related effects. Maturitas 2014; 79:428–434

38. Macaulay AJ, Stern RS, Stanton SL: Psychological aspects of 211 female patients attending a urodynamic unit. J Psychosom Res 1991; 35:1–10

39. Ahn KS, Hong HP, Kweon HJ, et al.: Correlation between overactive bladder syndrome and obsessive compulsive disorder in women. Korean J Fam Med 2016; 37:25–30

40. Bogner HR, O’Donnell AJ, de Vries HF, et al.: The temporal relationship between anxiety disorders and urinary incontinence among community-dwelling adults. J Anxiety Disord 2011; 25:203–208

41. Drummond LM, Boschen MJ, Cullimore J, et al.: Physical complications of severe, chronic obsessive-compulsive disorder: A comparison with general psychiatric inpatients. Gen Hosp Psychiatry 2012; 34:618–625

42. Viechtbauer W: Publication bias in meta-analysis: Prevention, assessment and adjustments. Psychometrika 2007; 72:269–271

43. Weissman MM, Gross R, Fyer A, et al.: Interstitial Cystitis and Panic Disorder: A Potential Genetic Syndrome. Arch Gen Psychiatry 2004; 61:273–279

